# Impact of Centralized Care at Aortic Centers on Aortic Dissection Outcomes: A 20-Year Analysis of U.S. Hospitals

**DOI:** 10.1101/2025.05.14.25327652

**Authors:** Patrick D Conroy, Besher Tolaymat, Alec Schubert, Bruce Tjaden, Kenji Minakata, Philip Batista, Marc L. Schermerhorn, Joseph V Lombardi

**Affiliations:** Department of Vascular and Endovascular Surgery, Cooper University Hospital, Camden, NJ; Department of Cardiothoracic Surgery, Cooper University Hospital, Camden, NJ; Department of Surgery, Division of Vascular and Endovascular Surgery, Beth Israel Deaconess Medical Center, Harvard Medical School, Boston, MA; Department of Vascular Surgery, AtlantiCare Regional Medical Center, Pomona, NJ

## Abstract

**BACKGROUND:** Over the past thirty years in the United States, hospitals have increasingly become incorporated into hospital systems, leading to organized care with more complex cases being managed at large urban-teaching hospitals. Over a similar period, changes in intervention guidelines for Aortic Dissection have occurred, with continually growing options for endovascular, minimally-invasive treatment. Given these dynamic changes, we examined trends in Aortic Dissection hospitalizations, intervention approaches, and hospital characteristics over past twenty-years to elucidate the effect of centralized care on outcomes.

**METHODS:** We identified all patients presenting with aortic dissection, both ascending and descending, (ICD-9-CM: 441.0; ICD-10-CM: I71.0) in the NIS between 2000-2021. We then examined the utilization of open repair, endovascular and complex-endovascular repair, as well as nonoperative/medical management. Stratified by hospital setting (urban-teaching, urban-nonteaching, and rural), we analyzed trends of interventions and in-hospital mortality over time. If an operation was performed, we were able to discern between ascending/arch or descending aorta after the 2017 ICD revision.

**RESULTS:** 553,030 patients with aortic dissection were identified. The number of inpatients in the US with aortic dissections has increased, with an incidence of 26.7 cases/100k in 2000 to 47.2 cases/100k in 2020 (p<0.01). Overall, including all hospital settings, aortic dissections were less frequently managed nonoperatively (2000-2021: 83%-71%) and more frequently managed endovascularly (Figure), with 85% of all descending and 16% of all ascending/arch aortic dissections undergoing TEVAR in 2021. Over time, aortic dissections have increasingly been managed at urban-teaching hospitals (2000-2021: 72%-92%;p<0.01). Since 2016, urban-teaching hospitals more frequently intervened on aortic dissections compared to their rural counterparts (21% vs. 6%;p<0.01), despite having similar rates of failed medical management (9.8% vs. 8.2%;p=0.30). Finally, comparing the last 5-years, urban-teaching hospitals have lower mortality rates when managing aortic dissection versus their rural counterparts (10.9% vs 11.7%, OR=1.10;p=0.02) and if managed operatively, there was a lower associated risk of mortality at urban-teaching hospitals compared to urban-nonteaching hospitals (12.5% vs. 17.3%, OR=1.46;p<0.01).

**CONCLUSION:** Both aortic dissection hospitalizations and interventions have significantly increased over the past two decades in the US. The growth of large hospital systems and their absorption of smaller hospitals into integrated primary through quaternary care centers has resulted in an increase in “regionalization” of care, in which complex cases are transferred to larger urban teaching centers. Our analysis suggests there is a mortality benefit from the centralization of aortic care to tertiary/urban-teaching centers, though further research into this question is required.

## Introduction

Aortic dissections (AD) continue to be a leading cause of morbidity and mortality amongst aortic pathologies. Acute dissections in particular remain highly catastrophic due to the risk of cardiac tamponade, visceral and limb ischemia, stroke, and rupture^1–3^. Therapy options for those with complicated and uncomplicated aortic dissections have always included conservative medical management and open surgical repair, but in 2012 management was revolutionized with the FDA approval of the first Thoracic Endovascular Aortic Repair (TEVAR) graft to treat AD^4^. Over the last decade, TEVAR has become the predominant modality of treatment for Type B Aortic Dissection (TBAD) in those meeting criteria for repair. Recently, TEVAR has even been used for repair of dissections involving the arch, ushering in the new complex endovascular era. Landmark trials, such as Study of Thoracic Aortic Type B Dissection Using Endoluminal Repair I and II (STABLE I and STABLE II), have provided evidence for favorable aortic remodeling and improved all-cause mortality after TEVAR at 1 and 5 years^5–9^. The advent and increased utilization of TEVAR has provided a mortality benefit, but there remain additional factors to consider when studying dissection outcomes.

Recent trend analyses have identified a number of factors contributing to the continual rise in prevalence of AD and persistent associated mortality risk. Patient factors such as tobacco use, obesity, uncontrolled hypertension, Caucasian race, advanced age, congestive heart failure (CHF), and substance abuse have been highlighted as contributors to the development and increased mortality associated with this condition^1,2^. Additionally, several database and single-center studies have reported increased in-hospital mortality rates when the operation occurred outside of high volume and specialized aortic surgical centers^3,10,11^.

Current literature does not elaborate on the mortality differences between higher versus lower volume hospitals, in urban and rural settings, or the relevance of centers being academic institutes. Given this, our study aims to explore and further elucidate these differences in care and ability to manage this complex aortic condition. Furthermore, our hope is to highlight the centralization of AD care amongst tertiary and quaternary aortic centers and the impact this has had on management and patient outcomes.

## Methods

### Data Source

We conducted a retrospective cohort analysis of admissions of patients with Aortic Dissection as well as open and endovascular repairs of Aortic Dissection using the National Inpatient Sample (NIS). The data were obtained from the NIS database from 2000 through 2021, representing almost 95% of all inpatient admissions in 48 of 50 U.S. states^12–14^. The Cooper University Hospital institutional review board approved the present study and waived the need for written informed consent owing to the retrospective and de-identified nature of the NIS database.

### Study Population

We first identified all patients within the NIS with a diagnosis code in any position for Aortic Dissection at any aortic level (ICD-9-CM: 441.0x; ICD-10-CM: I71.0) (irrespective of whether they underwent a prior aortic procedure, either open or endovascular, as there is no way to delineate using the NIS database). Due to the nature of ICD coding, there is no way to determine if an aortic dissection was acute or chronic, but a sensitivity analysis was performed comparing patients who had concomitant diagnoses of aortic dissection and thoracic aortic aneurysm to those without mention of aortic aneurysm (Supplemental Figure 1). From 2000 to quarter three of 2015, patients were identified using ICD-9-CM codes similar to prior NIS trend studies^15,16^. For quarter four of 2015 through 2021, the patients were identified using ICD-10-CM codes as described in Table 1 in the Data Supplement. Patients were further stratified based on what intervention approach they underwent (nonoperative, open, or endovascular).

**Table 1:**
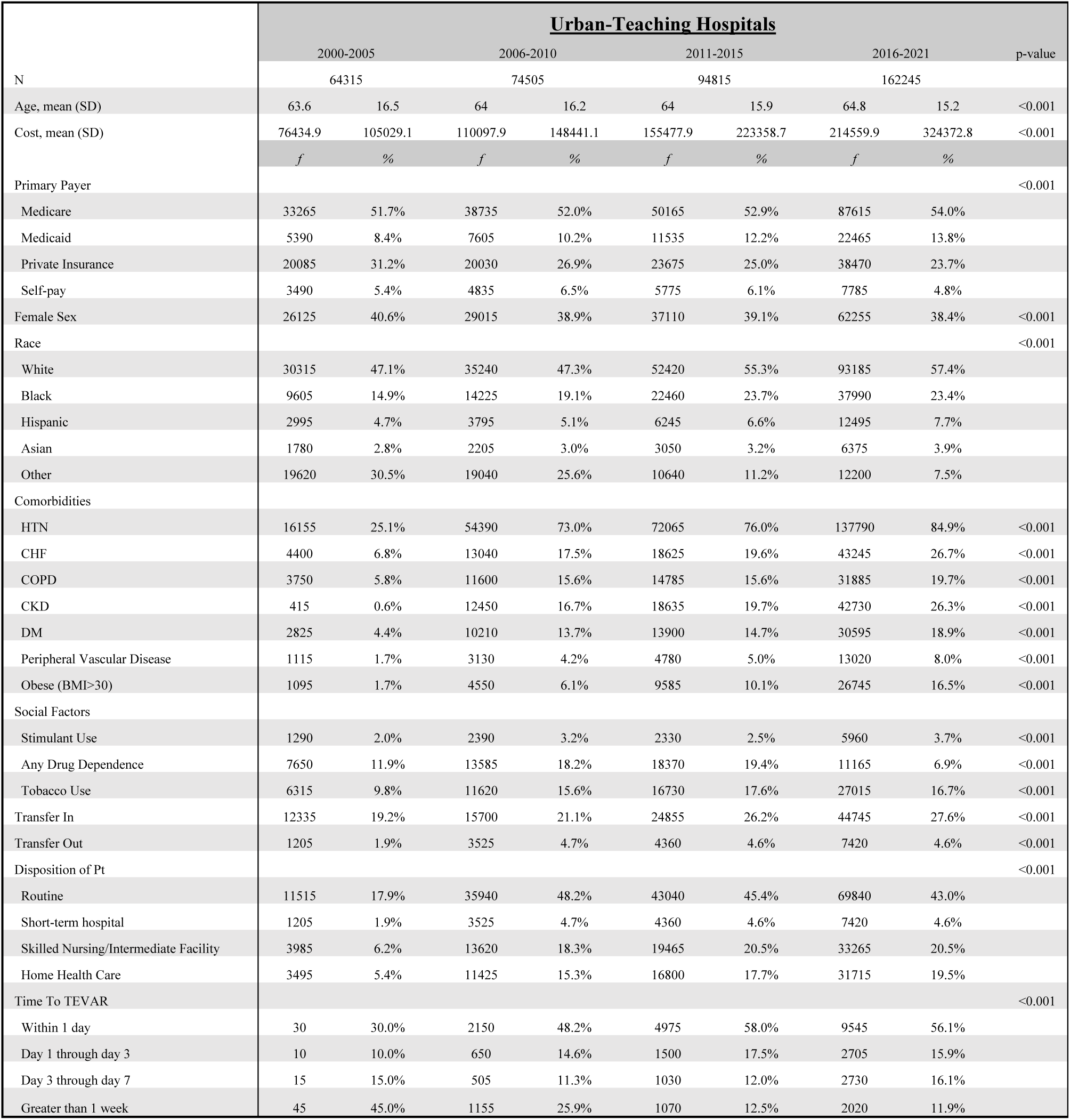
Demographics and comorbidities across the half-decades in Urban-Teaching Hospitals.

In order to ensure consistent data capture across ICD-9-CM to ICD-10-CM, we used validated methods to convert pathologies and procedures between coding systems^17–20^ (Supplemental Table 2).

### Study Definitions and Outcomes

Relevant comorbidity and demographic data were collected for all patients using Elixhauser covariates^21^ (2000-2012) and associated ICD codes (2013-2021). Our primary outcomes of interests were admissions, intervention approach, and in-hospital mortality. Intervention approaches for aortic dissection included in the study were nonoperative medical management, open aortic repair, and TEVAR, both simple and complex.

The nonoperative cohort included both patients who were trialed on medical therapy inpatient and discharged, as well as those who were transferred after trialing medical therapy. A sensitivity analysis was performed to determine the management of patients at “high transfer-rate hospitals,” with a threshold of transfer rate >10% of aortic dissection patients, to determine the effect of patients managed at low volume centers. (Supplemental Table 3).

The complex TEVAR (cTEVAR) cohort is based on ICD-10-CM codes capturing fenestrated or branched endografts (F/BEVAR), physician modified endografts (PMEG), and endovascular intervention utilizing the chimney/snorkel technique (chEVAR) within the thoracic aorta only. This intervention cohort does not represent patients with descending aortic dissections who underwent complex endovascular aortic repair within the visceral-segment alone.

More specific location of the dissection was able to be discerned after the 2017 revision of ICD-10-CM, when distinct procedure codes were produced for interventions either on the ascending thoracic aorta and arch or on the descending thoracic aorta. Prior to this revision, all thoracic pathologies were grouped together. Therefore, starting in 2017 we were able to discern between Ascending/Arch Thoracic Aortic Dissection and Descending Thoracic Aortic Dissection if patients underwent surgical repair during their hospitalization.

### Statistical Analysis

Continuous variables were represented as mean and standard deviation if they had normal Gaussian distribution while categorical variables were presented as frequencies and percentages. Continuous variables were compared using the Wald test to assess for differences in the mean while categorical variables were compared using the Pearson x^2^ test. Trends were analyzed with Cochrane-Armitage p-value trend test for presentations, interventions, and mortality over the years. Time-to-TEVAR trends were analyzed with Cuzick test for trend. Finally, for the last half-decade, from 2017-2021, we performed logistic regression comparing mortality after the different intervention approaches and in hospitals with different characteristics, while adjusting for key demographic and comorbidities. All statistical analyses were performed using Stata, version 17.0 (StataCorp, College Station, Texas).

## Results

### Overall Patient Demographics and Comorbidities (Table 1)

553,030 hospitalized patients were identified with Aortic Dissection in the total study period of 2000 to 2021. Stratified by hospital setting (Urban-Teaching – Table 1, Urban-Nonteaching – Table 2, Rural – Table 3), the demographics, comorbidities and transfer status were analyzed. Urban-Teaching hospitals was the only cohort to experience an increase in patient age (p-trend<0.001). Patients at all hospital settings experienced an increase in hospital-associated costs (all p-trend<0.001). All hospital settings saw an increased proportion of patients with Medicaid or Medicare over the past two decades (p-trend<0.001). Rural hospitals had the greatest number of Medicaid/Medicare patients (79%) in the last half decade, then Urban-Nonteaching hospitals (73%), then Urban-Teaching hospitals (68%).

**Table 2:** Demographics and comorbidities across the half-decades in Urban-Nonteaching Hospitals.

|  | Urban-Nonteaching Hospitals |  |  |  |  |  |  |  | p-value |
| --- | --- | --- | --- | --- | --- | --- | --- | --- | --- |
|  | 2000-2005 |  | 2006-2010 |  | 2011-2015 |  | 2016-2021 |  |  |
| N | 37740 |  | 39690 |  | 29950 |  | 21280 |  |  |
| Age, mean (SD) | 68.1 | 15.4 | 68.5 | 15.4 | 68 | 15.4 | 68.6 | 14.6 | 0.366 |
| Cost, mean (SD) | 52853.1 | 80500 | 81140.3 | 120931.2 | 111864.9 | 173175.7 | 144146.1 | 227876.9 | <0.001 |
|  | *f* | % | *f* | % | *f* | % | *f* | % |  |
| Primary Payer |  |  |  |  |  |  |  |  | <0.001 |
| Medicare | 23885 | 37.1% | 24970 | 33.5% | 18440 | 19.4% | 13380 | 8.2% |  |
| Medicaid | 2075 | 3.2% | 2690 | 3.6% | 2580 | 2.7% | 2220 | 1.4% |  |
| Private Insurance | 9300 | 14.5% | 9065 | 12.2% | 6230 | 6.6% | 4045 | 2.5% |  |
| Self-pay | 1445 | 2.2% | 1725 | 2.3% | 1565 | 1.7% | 890 | 0.5% |  |
| Female Sex | 16090 | 25.0% | 17450 | 23.4% | 12670 | 13.4% | 8770 | 5.4% | 0.061 |
| Race |  |  |  |  |  |  |  |  | <0.001 |
| White | 22455 | 34.9% | 24265 | 32.6% | 19930 | 21.0% | 13960 | 8.6% |  |
| Black | 3395 | 5.3% | 4775 | 6.4% | 4555 | 4.8% | 3490 | 2.2% |  |
| Hispanic | 2000 | 3.1% | 2180 | 2.9% | 2200 | 2.3% | 1715 | 1.1% |  |
| Asian | 715 | 1.1% | 1255 | 1.7% | 1160 | 1.2% | 910 | 0.6% |  |
| Other | 9175 | 14.3% | 7215 | 9.7% | 2105 | 2.2% | 1205 | 0.7% |  |
| Comorbidities |  |  |  |  |  |  |  |  |  |
| HTN | 9990 | 15.5% | 28935 | 38.8% | 22940 | 24.2% | 18095 | 11.2% | <0.001 |
| CHF | 3090 | 4.8% | 7920 | 10.6% | 6295 | 6.6% | 6215 | 3.8% | <0.001 |
| COPD | 3265 | 5.1% | 9290 | 12.5% | 6105 | 6.4% | 5275 | 3.3% | <0.001 |
| CKD | 350 | 0.5% | 7065 | 9.5% | 6555 | 6.9% | 6085 | 3.8% | <0.001 |
| DM | 1770 | 2.8% | 6540 | 8.8% | 5125 | 5.4% | 4540 | 2.8% | <0.001 |
| Peripheral Vascular Disease | 710 | 1.1% | 2055 | 2.8% | 1610 | 1.7% | 2080 | 1.3% | <0.001 |
| Obese (BMI>30) | 785 | 1.2% | 2890 | 3.9% | 2850 | 3.0% | 3305 | 2.0% | <0.001 |
| Social Factors |  |  |  |  |  |  |  |  |  |
| Stimulant Use | 450 | 0.7% | 645 | 0.9% | 515 | 0.5% | 645 | 0.4% | <0.001 |
| Any Drug Dependence | 4725 | 7.3% | 6415 | 8.6% | 5755 | 6.1% | 1210 | 0.7% | <0.001 |
| Tobacco Use | 4320 | 6.7% | 5825 | 7.8% | 5295 | 5.6% | 3315 | 2.0% | <0.001 |
| Transfer In | 2565 | 4.0% | 3535 | 4.7% | 2510 | 2.6% | 2060 | 1.3% | <0.001 |
| Transfer Out | 1860 | 2.9% | 4510 | 6.1% | 3435 | 3.6% | 2425 | 1.5% | <0.001 |
| Disposition of Pt |  |  |  |  |  |  |  |  | <0.001 |
| Routine | 6330 | 9.8% | 16920 | 22.7% | 12265 | 12.9% | 8410 | 5.2% |  |
| Short-term hospital | 1860 | 2.9% | 4510 | 6.1% | 3435 | 3.6% | 2425 | 1.5% |  |
| Skilled Nursing/Intermediate Facility | 2495 | 3.9% | 7605 | 10.2% | 6030 | 6.4% | 4185 | 2.6% |  |
| Home Health Care | 1840 | 2.9% | 5360 | 7.2% | 4455 | 4.7% | 3515 | 2.2% |  |
| Time To TEVAR |  |  |  |  |  |  |  |  | 0.706 |
| Within 1 day | 10 | 66.7% | 780 | 58.0% | 745 | 57.3% | 500 | 52.4% |  |
| Day 1 through day 3 | 0 | 0.0% | 205 | 15.2% | 175 | 13.5% | 170 | 17.8% |  |
| Day 3 through day 7 | 0 | 0.0% | 135 | 10.0% | 165 | 12.7% | 160 | 16.8% |  |
| Greater than 1 week | 5 | 33.3% | 225 | 16.7% | 215 | 16.5% | 125 | 13.0% |  |

**Table 3:**
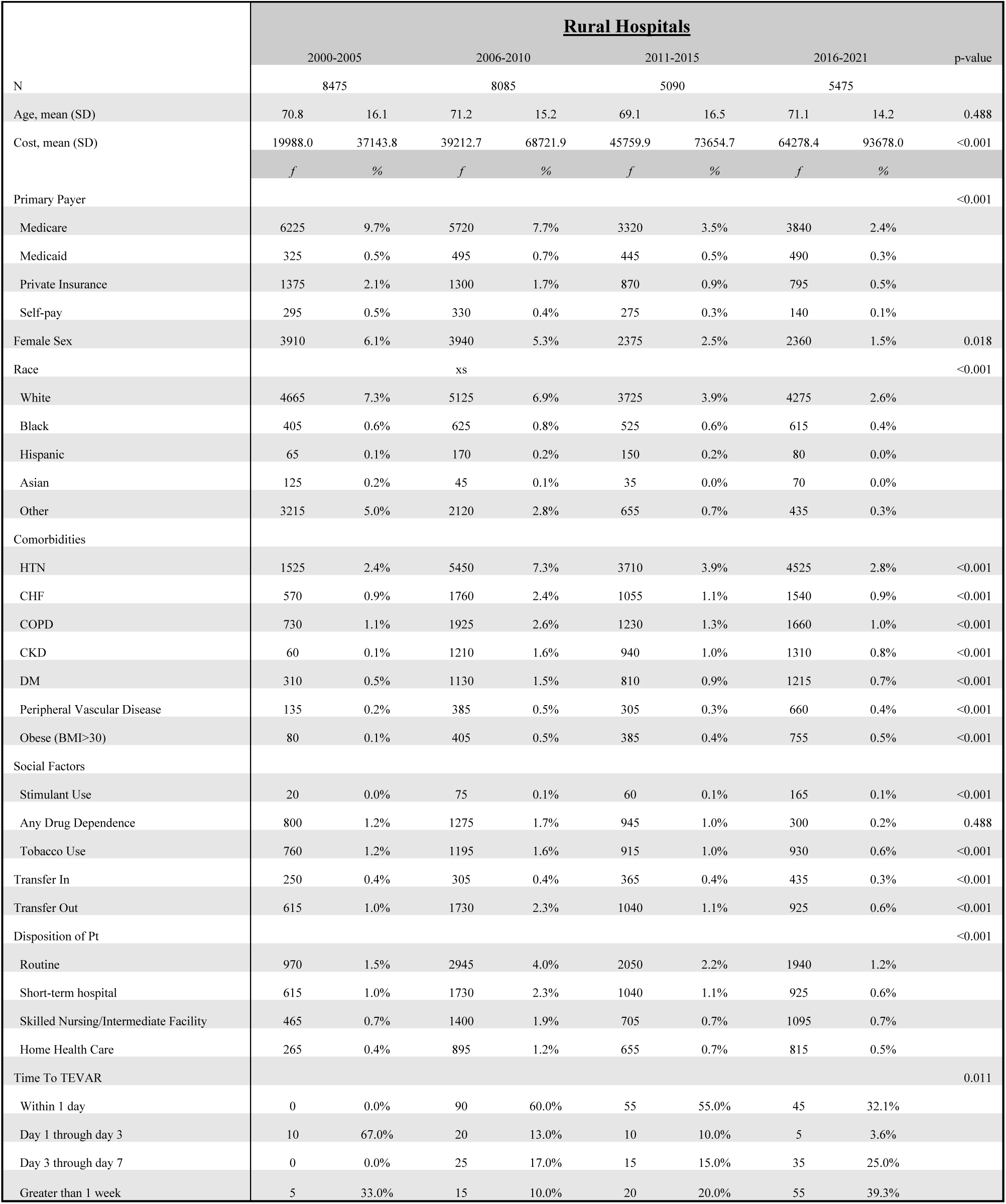
Demographics and comorbidities across the half-decades in Rural Hospitals.

**Table 4:**
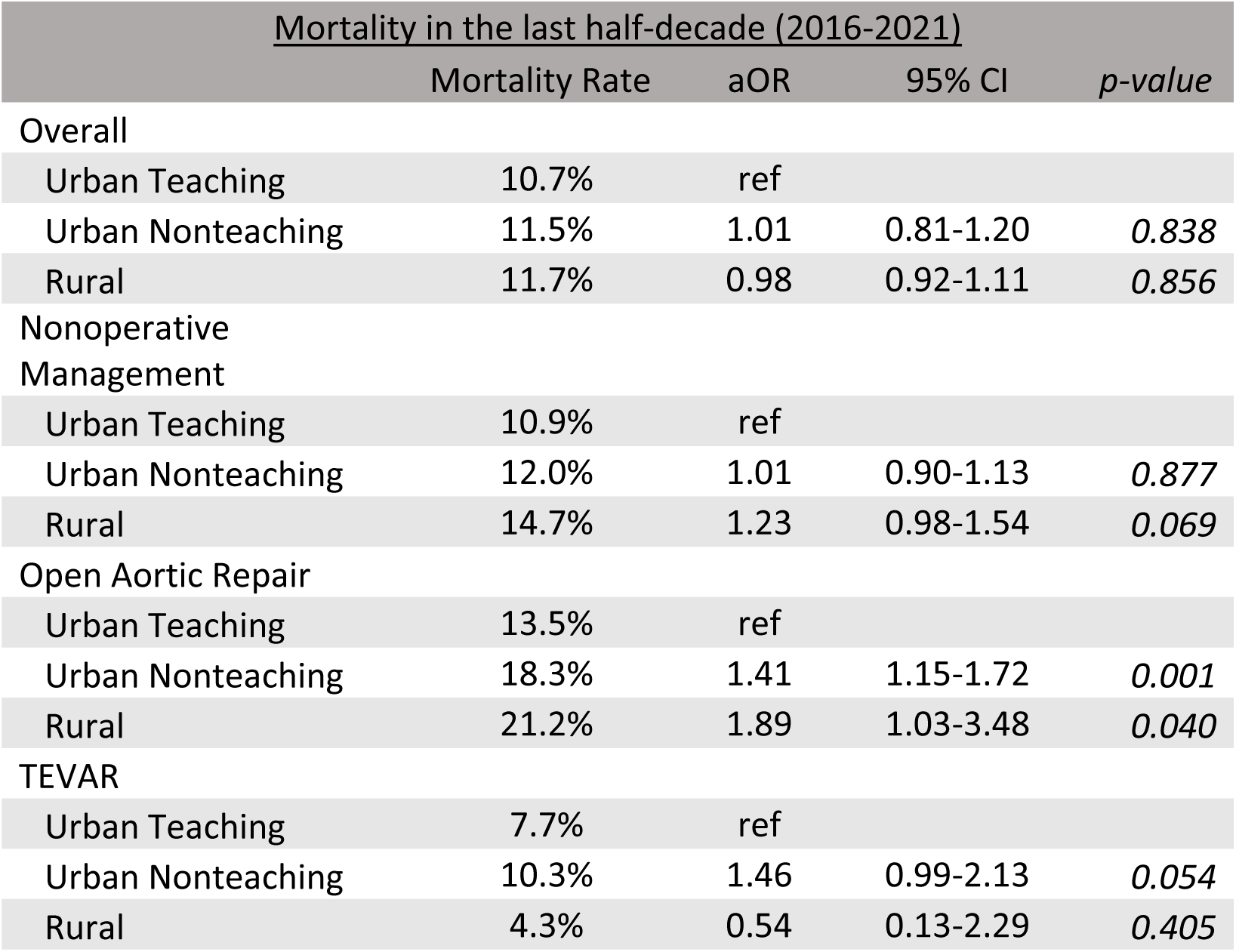
Logistic regression of outcomes stratified by hospital setting (Urban-Teaching, Urban-Nonteaching, Rural).

All hospitals saw similar proportions of female patients, accounting for approximately 40% of the overall AD population. Across all hospital settings, every hospital setting saw an increasing proportion of white patients over the years, with Urban-Teaching hospitals remaining the most racially diverse (White Race in 2021: Rural 83%, Urban-Nonteaching 61%, Urban-Teaching 58%).

All hospital settings treated patients with more reported comorbidities, with every comorbidity significantly increasing over the years. The most common comorbidity amongst AD patients was hypertension, present in 84% of all AD patients in the last 5 years. Also of note, AD patients were increasingly obese (2004 to 2021: 4.3% to 18.4%, p-trend<0.001) over the study period. Stimulant use increased amongst AD patients in all hospital settings, from 1.4% in 2000 to 3.9% in 2021 of all AD patients reporting non-prescribed stimulant use (p<0.001). Similar trends were seen in tobacco abuse amongst AD patients, with every hospital setting seeing nearly a doubling over the two-decade period (overall 2000 to 2021: 8.0%-15.3%; p<0.001).

AD patients in Urban-Teaching hospitals were increasingly transferred in (2000 to 2021: 18% to 27%). Conversely, AD patients in Urban-Nonteaching and Rural hospitals were increasingly transferred out (2000 to 2021: Urban-Nonteaching 0% to 12%, Rural 0% to 16%, both p-trend<0.001). All hospital settings saw similar trends of patients being discharged home ∼40% of the time.

Finally, we analyzed “Time-to-TEVAR” and the trends over time for each hospital setting. Urban-Teaching hospitals consistently demonstrated the shortest Time-to-TEVAR (2021: average 1.6 days), while rural hospitals had the longest Time-to-TEVAR (2021: average 3.9 days). Urban-Teaching hospitals saw a significant trend in more TEVARs being performed within 24 hours of presentation (49.6% in 2006 to 52.6% in 2021, p<0.001) while conversely, Rural hospitals saw a decrease (100% in 2006 to 27.2% in 2021, p<0.011). In the last half-decade, the majority of patients who received at TEVAR in Urban-Teaching and Urban-Nonteaching hospitals underwent their procedure within 1 day (56.1% and 52.4%, respectively), while 32.1% of patients in Rural hospitals underwent TEVAR within 1 day.

### Trends in Admissions and Repairs (Figure 1)

From 2000 to 2021, hospitalizations for AD increased significantly from 15,015 to 35,310 (p-trend<0.01). The prevalence of AD hospitalizations per 100,000 population doubled over the two decades, from 5.3 in 2000 to 10.6 in 2021 (Figure 1; p-trend<0.01). Out of the 553,030 total identified AD patients, 405,105 (73%) were treated nonoperatively, 103,425 (19%) underwent open repair, and 42,465 (8%) underwent endovascular repair. Less than 1% of all AD patients had a complex endovascular repair (2,035).

**Figure 1:**
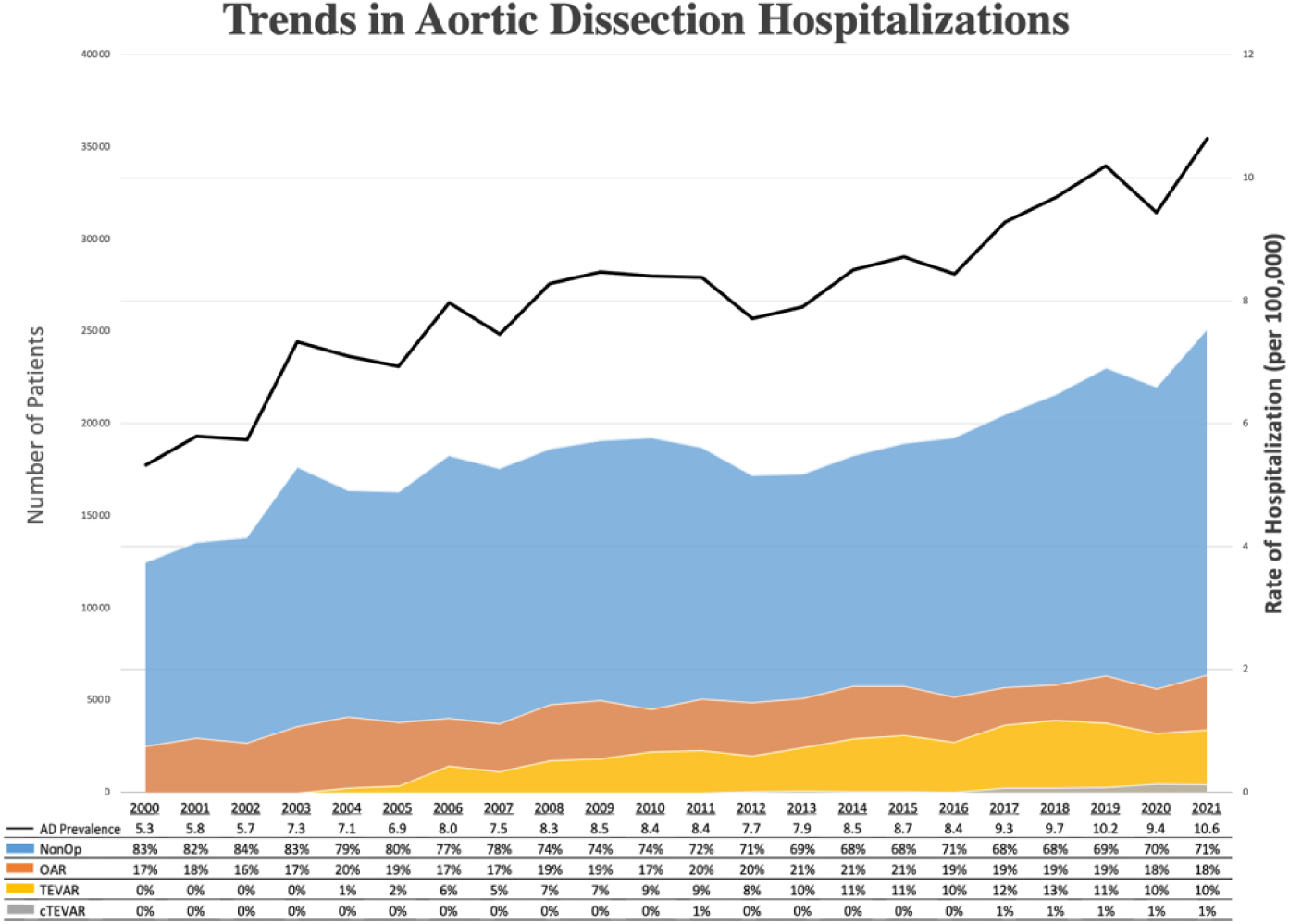
Aortic Dissection Hospitalization Prevalence with proportion of each intervention approach.

Over time there were minor shifts in management of AD. Despite remaining the most common approach, nonoperative management decreased from 83% in 2000 to 71% in 2021 (p-trend<0.01). Open aortic repair remained stable proportionally (2000-2021: 17%-18%; p-trend<0.01), with the absolute number of open repairs increasing from 2,525 to 6,360 during the same period. Thoracic endovascular aortic repair emerged as a treatment option in 2004, and its utilization increased substantially, with 10% of AD managed with TEVAR in 2021 (2005 to 2021: 365 to 3,410; p-trend<0.01). Complex endovascular repair was introduced in 2012, with 85 AD patients undergoing cTEVAR, and while the management became more common (435 in 2021), cTEVAR never accounted for more than 1% of all AD management. In 2021, cTEVAR accounted for 12.8% of all endovascular procedures done on aortic dissection.

### Trends in Admissions and Repairs by Hospital Setting (Figure 2)

Overall, 551,665 (99%) of AD patients identified had hospital characteristic data. Most AD patients (95%) were treated at Urban hospitals, while 5% were treated at Rural hospitals.

**Figure 2:**
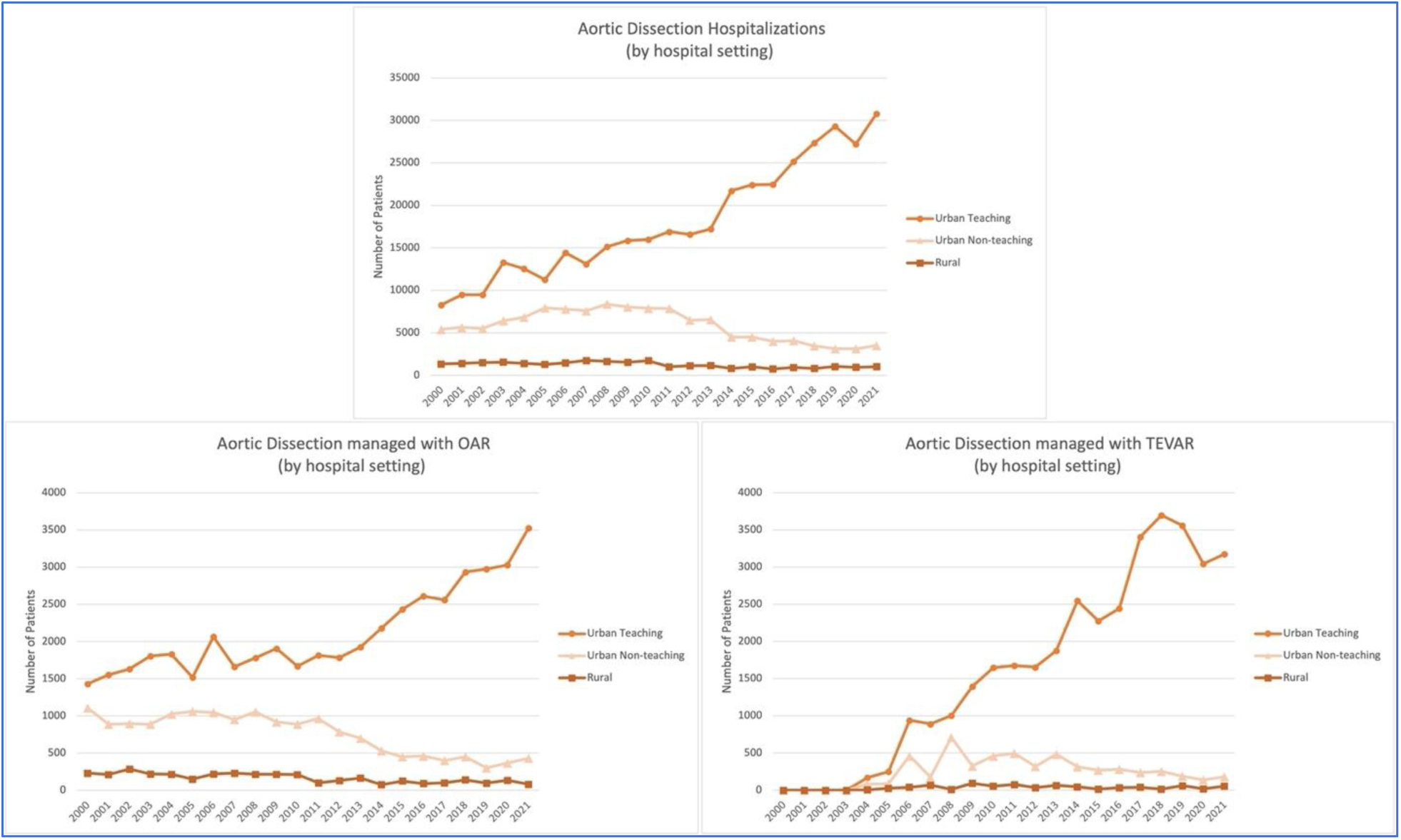
Aortic Dissection Hospitalizations and Intervention Approaches stratified based on hospital setting (urban teaching, urban non-teaching, rural) analyzing overall hospitalizations, patients who underwent OAR, patients who underwent TEVAR.

Urban-Teaching hospitals saw the highest volume of AD patients (72%), increasing significantly from 8,290 in 2000 to 30,790 in 2021 (p-trend<0.001; Figure 2). In contrast, Urban-Nonteaching and Rural hospitals saw a decreasing trend (2000 to 2021: Urban Nonteaching 5,395 to 3,505; Rural 1,330 to 1,015; both p-trend<0.001). In all years, nonoperative management remained the predominant approach across all hospital settings. Of the AD patients not transferred out in 2021, 92% were managed nonoperatively in Rural hospitals and 87% in Urban-Nonteaching hospitals. In Urban-Teaching hospitals, 71% of AD patients were managed nonoperatively in 2021.

Both open and endovascular repairs of AD increased dramatically in Urban-Teaching hospitals (2000 to 2021: OAR 1,430 to 3,525, p-trend<0.001) (2005 to 2021: TEVAR 250 to 3,175, p-trend<0.001), being performed with nearly equal frequency in recent years. In Urban-

Nonteaching hospitals, OAR reduced by more than half (2000 to 2021: 1,110 to 430; p-trend<0.001) while TEVAR was quickly adopted after 2005, with utilization peaking at 710 in 2008 (9% of all AD patients at Urban-Nonteaching hospitals), but then quickly down-trending to 180 TEVAR cases in 2021 (p-trend<0.001). In Rural hospitals, both open and endovascular repairs of AD remained the same (2000 to 2021: OAR 230 to 80, p-trend=0.096) (2005 to 2021: TEVAR 25 to 55, p-trend=0.153).

### Mortality Rate based on Intervention Approach stratified by Hospital Setting (Figure 3)

Over the study period, AD patients managed nonoperatively had a national mortality rate of 11.0%. Rural hospitals had the highest in-hospital mortality rate at 12.7%, followed by Urban-Nonteaching hospitals at 11.1%. Urban-Teaching hospitals demonstrated the lowest mortality rate after nonoperative management of AD, at 10.8%. In the nonoperative cohort, Urban-Teaching hospitals had significantly lower mortality rates when compared to either Urban-Nonteaching or Rural hospitals (both p<0.001). Similarly, Urban-Nonteaching hospitals had a significantly lower mortality rate than Rural hospitals (p<0.001; Figure 3).

**Figure 3:**
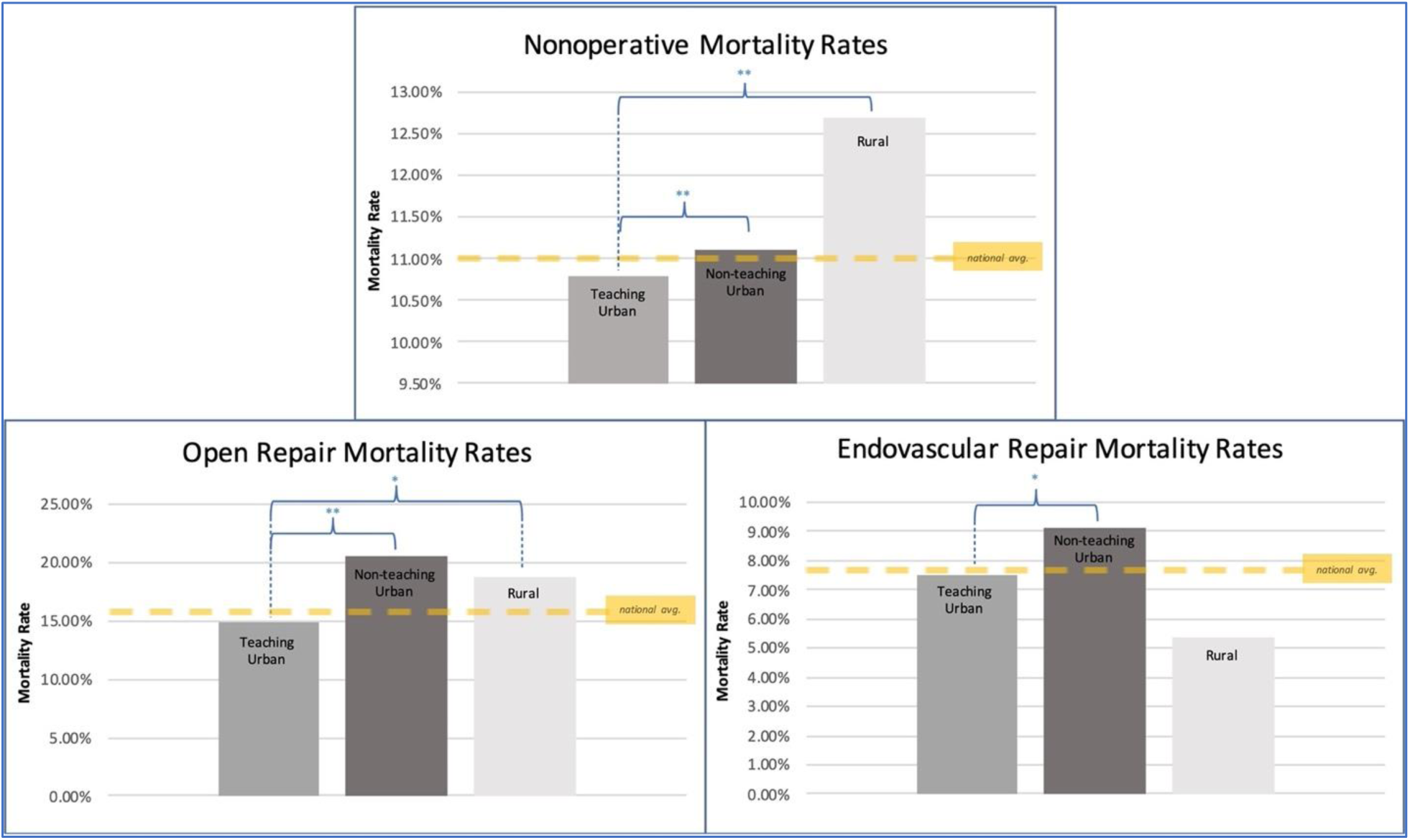
Mortality rate of aortic dissections by hospital setting (urban teaching, urban non-teaching, and rural) after nonoperative management, open repair, and endovascular repair (* is p<0.05; ** is p<0.0001).

For patients undergoing open repair over the study period, the national mortality rate was 15.7%. Rural hospitals had an in-hospital mortality rate at 18.8%, while Urban-Nonteaching hospitals had the highest mortality rate at 20.6%. Urban-Teaching hospitals again demonstrated the lowest mortality rate after open repair of AD, at 14.8%. In the OAR cohort, Urban-Teaching hospitals had significantly lower mortality rates when compared to either Urban-Nonteaching or Rural hospitals (p<0.001, p=0.037, respectively). There was no difference between in-hospital mortality rate after OAR at Urban-Nonteaching and Rural hospitals (p=0.572).

Over the entire study period, the national mortality rate after endovascular repair for AD was 7.7%. In the TEVAR cohort, Rural hospitals had the lowest mortality rate at 5.4%, though there were less than 800 TEVARs done at Rural hospitals over the past 15 years. Urban-Nonteaching hospitals again had the highest mortality rate at 9.1%, while Urban-Teaching hospitals demonstrated an in-hospital mortality rate after TEVAR at 7.5%, with the majority of TEVARs being performed in this cohort. Urban-Teaching hospitals had significantly lower mortality rates when compared to Urban-Nonteaching hospitals (p=0.042). There was no difference between Rural hospitals and either Urban cohort, though this is most likely due to small sample size in that cohort with only 760 TEVARs done at Rural hospitals between 2004 and 2021 (Rural:Urban-Nonteaching p=0.055; Rural:Urban-Teaching p=0.176).

### Trends in Thoracic Aorta Ascending/Arch vs. Descending Subanalysis (Figure 4)

To further stratify intervention approach cohorts into Arch or Ascending Thoracic Repair versus Descending Thoracic Aortic Repair, 45,845 patients with granular data were identified between 2017 and 2021. A majority of the 30,675 Arch/Ascending Thoracic AD included were repaired open: 86% underwent OAR and 14% underwent TEVAR. In contrast, a majority of the repairs to the 15,170 Descending Thoracic AD were performed endovascularly (82% TEVAR, 18% OAR)

**Figure 4:**
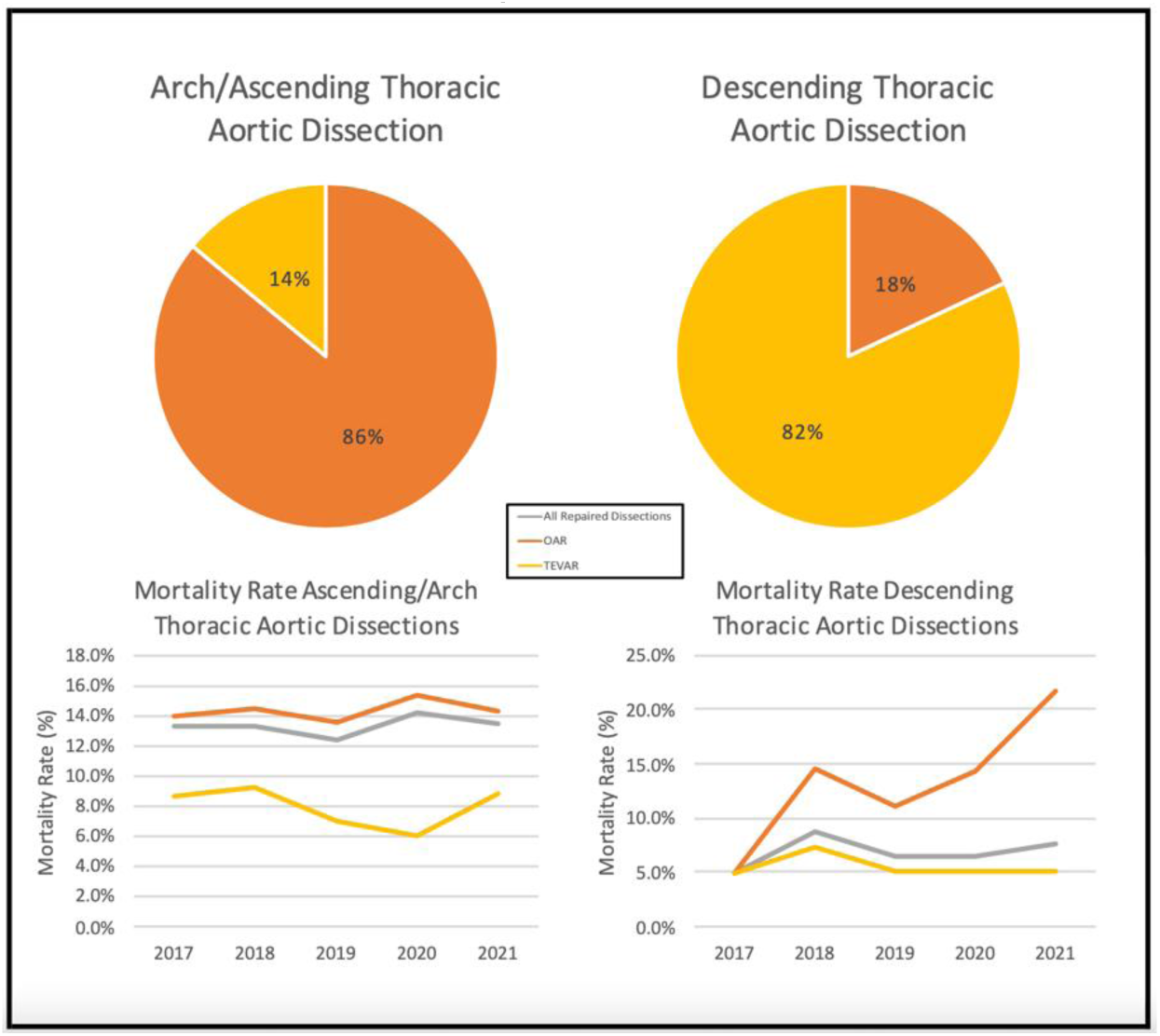
Subgroup analysis of intervention approach and mortality rate of separated Arch/Ascending Thoracic versus Descending Thoracic Aortic Dissections.

The overall mortality rate after any repair of Arch/Ascending Thoracic AD fluctuated around 13.5% over the five years (p-trend=0.544), with a stable mortality rate after TEVAR averaging 7.0% (p-trend=0.987) and after OAR averaging 14.3% (p-trend=0.975). Conversely, the overall mortality rate after any repair of Descending Thoracic AD was stable and averaged 6.8% over the past five years (p=0.110), with a stable mortality rate after TEVAR averaging 5.5% (p-trend=0.705) and substantially increasing in-hospital mortality rate after OAR (2017 to 2021: 5.0% to 21.7%, p<0.001).

### Logistic Regression Analysis of Mortality Rates between Hospital Settings (Table 2)

Data was analyzed from the past five years in order to reflect contemporary practice trends after the ICD-10 revision. During the last half-decade, overall mortality rates were similar across hospital types in the logistic regression model. Adjusted for important demographics and comorbidities, with Urban-Teaching hospitals as reference, Urban-Nonteaching hospitals had no significant difference in mortality rate (aOR=1.01, p=0.84), nor did Rural hospitals (aOR=0.98, p=086). For patients managed nonoperatively, there was no difference in mortality between Urban-Nonteaching and Urban-Teaching hospitals (aOR=1.01, p=0.88), while Rural hospitals tended to be associated with higher rates of mortality, though this trend did not reach statistical significance (aOR=1.23, p=0.07).

Patients who underwent OAR at Urban-Nonteaching hospitals had a higher associated mortality rate when compared with those who underwent OAR at Urban-Teaching hospitals (aOR=1.41, p<0.01). Similarly, patients who underwent OAR at Rural hospitals also had a higher associated mortality rate (aOR=1.89, p=0.04). Finally, patients who underwent TEVAR at Urban-Nonteaching hospitals trended to have higher associated mortality rate (aOR=1.46, 0.05) while Rural hospitals saw no difference from their Urban-Teaching counterparts (aOR=0.54, p=0.41).

## Discussion

In this nationwide retrospective study spanning two decades, we present evidence supporting the centralization of care at aortic centers in the management of Aortic Dissection.

Academic centers that act as tertiary or quaternary hospitals, often at the center of larger referral networks, produce better outcomes managing Aortic Dissections, even when patients are managed nonoperatively. Robust transfer systems, therefore, are paramount to ensuring that patients with AD can be cared for with the highest level of care, minimizing morbidity and mortality. This becomes even more important in this era when the prevalence of AD continues to increase in the United States, nearly doubling over the twenty-year study period from 5.3 to 10.3 per 100,000.

Modern, guideline-based management of aortic dissection, acute or chronic, is dynamic and requires multimodal care by experienced teams. Whether the patient is managed medically or surgically, AD patients are resource intensive, require multiple specialties to be available, and will need to be followed lifelong to monitor for disease progression, as aneurysmal degeneration and re-interventions are not uncommon. Medical management requires initial admission to the intensive care unit, impulse control with intravenous antihypertensives, and close physiologic monitoring of factors that may necessitate a shift to operative management, such as frequent neurologic and pulse exams with pain assessments, as well as monitoring for evidence of malperfusion with a trained and nuanced eye^22^. The highly morbid complications of dissection require prompt diagnosis and expeditious repair, and the post-operative period carries its own unique and resource intensive considerations (e.g. the need for neurologic monitoring and the availability of neurosurgery for emergent lumbar drain placement if spinal cord ischemia occurs).

We found disparities in mortality rates between Rural hospitals and their larger academic counterparts, potentially demonstrating larger hospital’s ability to provide better care for AD patients given more extensive resources and the immediate availability of multiple medical and surgical specialties. Rural hospitals had significantly higher mortality rates for AD patients managed nonoperatively (18% relative increase) and with open repair (27% relative increase) compared with Urban-Teaching centers. Our reported disparities are concordant with prior literature that have described higher mortality rates of Type A AD patients admitted to smaller hospitals^23^ and higher rates of initial misdiagnosis in non-teaching centers^24^. Similarly, when analyzing aortic aneurysms, Ahuja et al. reported higher mortality rates in rural settings compared to their urban counterparts^25^ and multiple studies have even linked higher rates of mortality and readmission proportional to greater distances from tertiary care hospitals^26,27^.

We also present a significant increase – more than tripling – in the volume of admitted AD patients in the 21-year duration of the study, being the first to span ICD-9 and ICD-10 with code conversion. This increase is likely multifactorial in nature, both highlighting worsening health of the U.S. population and improvements in hospital diagnostic modalities. Hypertension is a known leading cause of dissection, is highly co-incident in our cohort, and has been described to be increasing in incidence nationally from 1999 to 2018^28^. The aging population in the United States has similarly increased significantly, with the largest increase of the 65+ year old population occurring from 2010-2020^29^. Simultaneously, the rate of diagnosing aortic dissection is likely to have increased during this period due to the increased use and more-readily available CT scans, which have increased in utility by 90% in the past decade in a recent study of Medicare data^30^.

With increasing incidence of aortic pathologies^31^ and evidence of disparity of outcomes between different types of hospitals, interest in centralization of specialized care has become an increasingly discussed topic around the globe^32,33^. Countries like England and Finland have pushed for centralization of care, and many argue this has led to tangible improvements in overall national outcomes for aortic care^34–36^. Since the 1990s, the overall U.S. healthcare system has similarly observed a trend towards centralization of specialized care with hospital mergers and system consolidations. Initially these consolidation efforts were likely made with financial considerations at the forefront^37–39^, and there remains debate as to whether this has conferred a benefit on patient outcomes or experience^40,41^. Despite questions as to the effects of hospital consolidation to overall patient care, there have been numerous studies that have demonstrated outcome benefits to consolidation of surgical care to high-volume centers in cardiac^42,43^ and vascular^44^ procedures. Our study adds to this body of work by highlighting the national benefits of AD management at high-volume, academic institutions, particularly in conferring mortality benefits regardless of management approach.

Limitations for this investigation are both due to the nature of the NIS database used, as well as the chronicity and readmission rates associated with AD. While the NIS is a robust database, accounting for 20% of all inpatient admissions in the U.S., it is susceptible to documentation and coding errors. This has been investigated in the past with extensive internal and external validation^45,46^ . Also, there is potential for selection bias given that this is an observation analysis. Finally, a subset of the captured admissions are likely ‘double counted’ as (1) re-admissions of the same patient, especially as re-admission has been previously shown to be common occurrence at a rate of ∼30% at 90 days following repair^47,48^, as well as (2) transfers from one hospital to another. Given the current paradigm of management, patients often undergo urgent open repair of their type A dissection and will predictably require re-admissions for management of the residual type B dissection.

## Conclusion

High-quality care of aortic dissection is resource-intensive, dynamic, multi-disciplinary, and timely – requirements that are best served at tertiary care centers and often, teaching hospitals. This model of dissection care as outlined is supported by our finding that 28% of patients were transferred <u>in</u> to Urban-Teaching hospitals whereas 10-17% were transferred <u>out</u> of Urban-Nonteaching and Rural hospitals. Assuming this subset of patients were transferred for higher level of care and therefore reflect a higher burden of disease, the lower mortality rate of the Urban-Teaching hospital setting is even more impactful and impressive. Considerations should be made in future guidelines not just on when to treat aortic dissection patients operatively, but also when to transfer patients to higher levels of care in a hope to improve overall aortic dissection outcomes.

## Data Availability

The National Inpatient Sample (NIS) database is national data supplied for a fee from the HCUP.

## Acknowledgments

None.

## Sources of Funding

This study had no funding sources.

## Disclosures

There are no financial conflicts of interest to disclose.

## Notes

### Competing Interest Statement

The authors have declared no competing interest.

### Funding Statement

No external funding was recieved.

### Author Declarations

Our institutional IRB approved this study, given it is a de-identified database.

